# SCORES (Selection of Clinical Observations for Risk Evaluation Scores): Impact of Heart Rate Selection on Simplified Geneva Score Performance in Suspected Pulmonary Embolism

**DOI:** 10.64898/2026.10.08.26365100

**Authors:** C.J. Cole

## Abstract

**Background:** The Simplified Geneva Score (SGS) incorporates two heart-rate thresholds. Multiple heart-rate measurements are commonly recorded during emergency department (ED) assessment. While the original SGS specifies use of mean heart rate, real-world implementation is unlikely to reflect this, and the effect of alternative heart-rate selection methods on score performance is unknown.

**Methods:** SCORES was a retrospective within-subject study of consecutive ED patients at a single urban hospital undergoing computed tomography pulmonary angiography (CTPA) for suspected pulmonary embolism (PE). Four SGS variants were calculated using the first, highest, mean, and lowest recorded pre-CTPA heart rate. Outcomes included score output, risk classification, diagnostic performance, reclassification flow, and downstream imaging utilisation. Precision estimates and comparisons were derived using empirical resampling techniques.

**Results:** 590 episodes were reviewed with 581 included for analysis. PE was confirmed in 50 cases (8.6%). Mean SGS differed significantly across all heart-rate implementations (p<0.0001). The proportion classified as high risk varied from 15% using the lowest heart rate to 40% using the highest heart rate. Reclassification across the SGS threshold was common, with 25% of episodes changing risk category unidirectionally between lowest and highest heart rate implementations. SGS(lowest HR) compared with SGS(mean HR) and SGS(highest HR) demonstrated higher specificity (87.6% vs 73.4% vs 62.1%), improved AUROC (0.744 vs 0.704 vs 0.659), and reduced downstream CTPA utilisation (818 vs 876 vs 920 scans per 100 PEs diagnosed), with no additional pulmonary emboli missed within the complete diagnostic pathway.

**Conclusions:** Heart-rate selection materially influences SGS output, risk classification, diagnostic performance, and imaging utilisation. In this retrospective cohort, use of the lowest recorded heart rate produced the most favourable overall balance of discrimination and diagnostic efficiency, supporting prospective validation.

## 1.0 Introduction

### 1.1 Background

Pulmonary embolism (PE) is a common and potentially life-threatening condition frequently investigated in emergency departments (EDs). Since the introduction of computed tomography pulmonary angiography (CTPA), imaging utilisation for suspected PE has increased substantially, without a concomitant increase in PE mortality (1), raising concerns regarding over-investigation, radiation exposure, contrast exposure, and healthcare resource utilisation.

To improve diagnostic efficiency, risk stratification tools (RSTs) such as the Wells score (2,3) and the Revised Geneva Score (RGS) and Simplified Geneva Score (SGS) (4,5) are commonly incorporated into diagnostic pathways. These tools assist clinicians in identifying patients with sufficiently low probability of PE to justify further assessment with downstream rule-out strategies, including PERC and D-dimer (XDP) testing, rather than immediate CTPA.

The SGS (Table 1) incorporates heart rate as two dichotomous variables (>74 bpm and >94 bpm). Although the original derivation and validation studies specified use of the mean heart rate when multiple measurements are available (4), this detail is not prominently described and is unlikely to reflect routine bedside practice. During a typical emergency department presentation, multiple heart rate values are usually recorded and clinicians may instead utilise the first recorded, highest observed, or most readily apparent value.

**Table 1.** The Simplified Geneva Score (SGS) for stratifying risk of pulmonary embolism.

| **Simplified Geneva Score** |  | < 3 = low-risk | > 2 = high-risk |
| --- | --- | --- | --- |
| **Clinical Feature** | **Points** |  |  |
| Age > 64 | 1 |  |  |
| Previous VTE | 1 |  |  |
| Surgery / major trauma < 1 month ago | 1 |  |  |
| Active malignancy | 1 |  |  |
| Haemoptysis | 1 |  |  |
| Unilateral lower limb pain | 1 |  |  |
| Unilateral lower limb oedema & tenderness | 1 |  |  |
| HR > 74 bpm | 1 |  |  |
| HR > 94 bpm | 1 |  |  |

#### 1.2 Knowledge gap

Whether the choice of heart rate value used in risk stratification tools significantly influences score output and diagnostic performance remains poorly characterised. Furthermore, the clinical relevance of any such differences is unknown. Given the role of these tools in guiding investigation and management, even modest differences may have implications for resource utilisation and patient safety.

#### 1.3 Objective

To determine whether the choice of heart rate value produces clinically meaningful differences in Simplified Geneva Score outputs, risk classification, and diagnostic performance in a real-world ED patient population.

### 2.0 Methods

#### 2.1 Ethics

Ethics approval was granted by the ACT Human Research Ethics Committee (HREC) as low-risk, with waiver of consent: 2025/ETH02245. The study was conducted in accordance with the National Statement on Ethical Conduct in Human Research (Australia), and the Declaration of Helsinki.

#### 2.2 Study design

SCORES was a retrospective within-subject score-performance benchmarking study comparing alternative implementations of the Simplified Geneva Score, differing only in their choice of heart rate value.

#### 2.3 Participants

Participants were consecutive ED patients undergoing CTPA for investigation of suspected PE during the study period (34 months from November 2022 to September 2025), at a single urban public hospital mixed (adult and paediatric) ED. A list of all instances of a CTPA being performed for a patient while in the ED during the study period was obtained from the Digital Health Record (DHR) system. Manual chart review was then undertaken. Cases with incomplete clinical data, or equivocal CTPA findings, were excluded. Analysis was per episode, rather than per patient.

#### 2.4 Patient and public involvement statement

SCORES was a retrospective observational study involving secondary use of de-identified extant clinical data, with a waiver of consent approved by the ACT HREC. There was no reasonable role or scope for patient or public involvement in the design or conduct of the study. The results will be disseminated in local forums and published via widely accessible platforms including medRxiv.

#### 2.5 Data management and security

Data management and security were compliant with local institutional and HREC requirements. Data entry was performed in a relational database using Microsoft Access. The master dataset was strongly encrypted, held on non-networked local media, and accessible only to the principal investigator. A de-identified working dataset, linked only by unique study identifier per record, was used for all analysis and reporting.

#### 2.6 Calculation of SGS from data

The Simplified Geneva Score (SGS) was chosen for analysis, as it is inherently more objective and less prone to inter-rater variability than the older Wells score for PE (4), and is more reliably calculated from retrospective examination of medical notes, minimising exclusion of cases due to incomplete data. The SGS and its dichotomous classification cut-off are shown in Table 1.

For each episode, manual DHR chart review was undertaken to extract the seven non-heart-rate features of the SGS, along with multiple heart rate measurements recorded during that episode of care in the ED, to permit reconstruction of the remaining two SGS components related to heart rate (HR > 74 bpm, HR > 94 bpm) using alternative heart rate values. Only heart rate values recorded prior to the ordering of the CTPA scan were included, reflecting the limitations of information available to the clinician at the time. The CTPA result (PE vs no PE) was also extracted.

For each episode, four SGS variants were then calculated based on different choices of heart rate (HR) value: first recorded HR, highest HR, lowest HR, and mean HR.

#### 2.7 Outcome measures

Outcome measures comprised score output, dichotomous risk classification, and diagnostic performance. Diagnostic performance was assessed using sensitivity, specificity, positive predictive value (PPV), negative predictive value (NPV), and area under the receiver operating characteristic curve (AUROC).

Reclassification flow between SGS variants was evaluated by quantifying movement across the SGS dichotomous classification threshold and the resulting impact on CTPA utilisation and number-needed-to-scan (NNS) at isolated tool and pathway levels. Pathway-level NNS was derived by applying each SGS implementation within the complete diagnostic pathway, using observed cohort data for downstream PERC and/or XDP assessment of low-risk patients to calculate the resulting number of CTPA examinations performed per PE diagnosed.

#### 2.8 Statistical analysis & software

Formatting, cleaning, and analysis of de-identified data were performed with Nim 2.2.8, transpiled to C, and compiled by Clang 16.0.0 on macOS 15.

Given the paired nature of the comparisons, discrete score outputs, non-Gaussian sampling distributions, and the absence of closed-form solutions for several derived performance metrics, empirical non-parametric methods were employed in preference to asymptotic approximations. Resampling was conducted via stratified bootstrapping to preserve baseline event prevalence across iterations, using 100,000 bootstrap replicates to maximise estimate stability. Highest Density Intervals (HDIs) were constructed for precision estimation, and empirical p-values were derived directly from the corresponding bootstrap distributions.

### 3.0 Results

#### 3.1 Cohort selection & characteristics

Query of the digital health record (DHR) system identified 3,554 episodes during the study period, in which CTPA was performed while the patient was in the ED. A total of 590 consecutive episodes underwent manual chart review, with 9 excluded due to incomplete data, yielding a final analysis dataset of 581 episodes. PE was confirmed in 50 cases (prevalence 8.6%). Patient age ranged from 15 to 99 years (mean 58 years, SD 19 years), and 56% of cases were female. Cohort characteristics are summarised in Table 2.

**Table 2.**
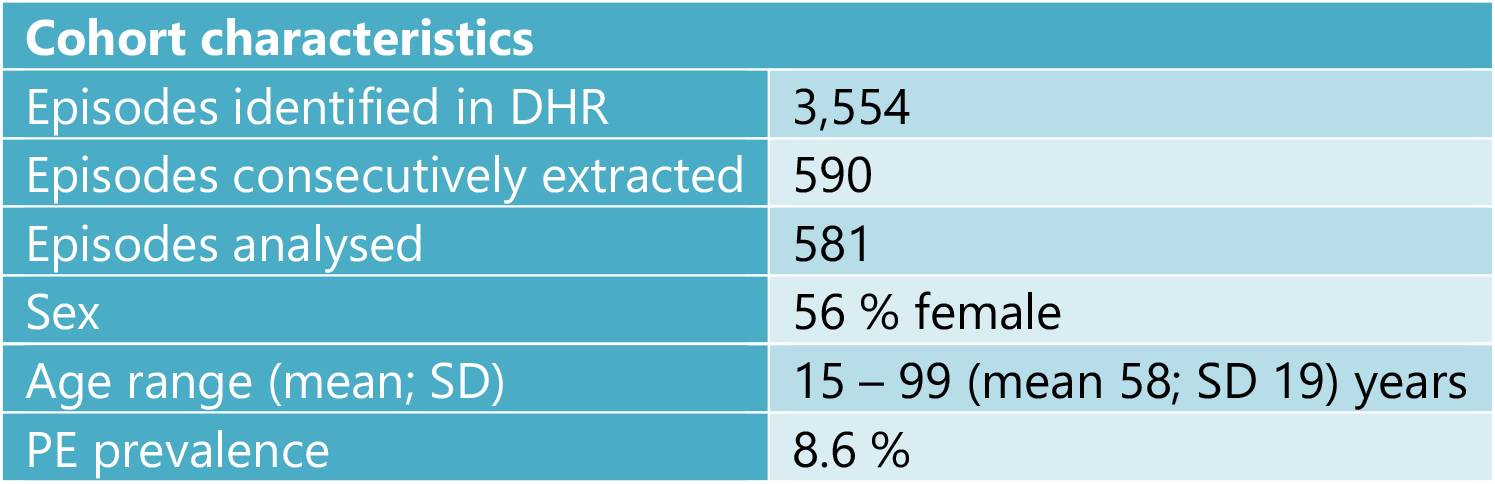
Cohort characteristics.

| Cohort characteristics |  |
| --- | --- |
| Episodes identified in DHR | 3,554 |
| Episodes consecutively extracted | 590 |
| Episodes analysed | 581 |
| Sex | 56 % female |
| Age range (mean; SD) | 15 – 99 (mean 58; SD 19) years |
| PE prevalence | 8.6 % |

### 3.1 Outcome metrics

#### 3.2.1 SGS output and risk classification

The four SGS variants were evaluated for score output and resulting dichotomous risk stratification. Mean SGS differed significantly (p < 0.0001) across all pairwise comparisons (Table 3).

**Table 3.** Diagnostic performance of SGS variants.

| SGS Variant | Mean SGS | AUROC | Sens. % | Spec. % | Proportion classed as high-risk | High-risk PE % (PPV) | Low-risk PE % (1-NPV) | NNS (tool) | NNS (pathway) |
| --- | --- | --- | --- | --- | --- | --- | --- | --- | --- |
| **Highest HR**<br>(95% HDI CI) | **2.31**<br>2.23-2.39 | **0.659**<br>0.581-0.737 | **66.0**<br>52.0-78.0 | **62.1**<br>58.0-66.1 | **40 %** | **14.1**<br>11.3-16.8 | **4.89**<br>3.10-6.74 | **4.7** | **9.2** |
| **First HR**<br>(95% HDI CI) | **2.12**<br>2.03-2.20 | **0.687**<br>0.612-0.763 | **64.0**<br>50.0-76.0 | **68.0**<br>64.2-71.9 | **35 %** | **15.8**<br>12.7-19.1 | **4.74**<br>3.11-6.45 | **4.0** | **9.0** |
| **Mean HR**<br>(95% HDI CI) | **1.95**<br>1.86-2.03 | **0.704**<br>0.629-0.776 | **60.0**<br>46.0-72.0 | **73.4**<br>69.7-77.0 | **29 %** | **17.5**<br>13.8-21.5 | **4.87**<br>3.33-6.49 | **3.4** | **8.8** |
| **Lowest HR**<br>(95% HDI CI) | **1.43**<br>1.34-1.51 | **0.744**<br>0.680-0.804 | **44.0**<br>32.0-58.0 | **87.6**<br>84.7-90.2 | **15 %** | **25.0**<br>18.1-32.6 | **5.67**<br>4.30-6.94 | **1.8** | **8.2** |

The proportion of cases classified as high risk ranged from 15% for SGS(lowest HR) to 40% for SGS(highest HR) across variants (Table 2). Significant differences in high-risk classification were observed between all SGS variants except SGS(first HR) and SGS(mean HR) (Table 4).

**Table 4.** Differences in output metrics: pairwise comparison of the 4 SGS variants. (Significant differences shown in **bold** typeface).

| Comparison | ΔAUROC | ΔSens. | ΔSpec. | ΔProportion high-risk | ΔHigh-risk PE % (ΔPPV) | ΔLow-risk PE % ($\Delta$(1-NPV)) |
| --- | --- | --- | --- | --- | --- | --- |
| Lowest vs Highest HR | **+ 0.085**<br>p = 0.00002 | **-22 %**<br>p < 0.00001 | **+ 25.5 %**<br>p < 0.00001 | **- 25 %**<br>p < 0.00001 | **+ 10.9 %**<br>p = 0.00002 | **+ 0.78 %**<br>p = 0.00002 |
| Lowest vs First HR | **+ 0.057**<br>p = 0.00294 | **-20 %**<br>p = 0.00002 | **+ 19.6 %**<br>p < 0.00001 | **- 20 %**<br>p < 0.00001 | **+ 9.2 %**<br>p = 0.00294 | **+ 0.93 %**<br>p = 0.00294 |
| Lowest vs Mean HR | **+ 0.040**<br>p = 0.02706 | **-16 %**<br>p = 0.00036 | **+ 14.2 %**<br>p < 0.00001 | **- 14 %**<br>p < 0.00001 | **+ 7.5 %**<br>p = 0.02706 | **+ 0.80 %**<br>p = 0.02706 |
| Mean vs Highest HR | **+ 0.045**<br>p = 0.00388 | - 6 %<br>p = 0.09074 | **+ 11.3 %**<br>p < 0.00001 | **- 11 %**<br>p < 0.00001 | **+ 3.4 %**<br>p = 0.00388 | **- 0.02 %**<br>p = 0.00388 |
| First vs Highest HR | **+ 0.028**<br>p = 0.01008 | - 2 %<br>p = 0.72916 | **+ 5.9 %**<br>p < 0.00001 | **- 5 %**<br>p < 0.00001 | **+ 1.7 %**<br>p = 0.01008 | **- 0.14 %**<br>p = 0.01008 |
| Mean vs First HR | + 0.017<br>p = 0.33256 | - 4 %<br>p = 0.44768 | **+ 5.4 %**<br>p = 0.00002 | **- 6 %**<br>p = 0.00002 | + 1.7 %<br>p = 0.33256 | + 0.13 %<br>p = 0.33256 |

#### 3.2.2 Diagnostic performance

Sensitivity varied markedly from 44 to 66 %, while specificity ranged from 62 to 88 %. Positive and negative predictive values showed corresponding variation across SGS variants (Table 3).

AUROC values ranged from 0.659 to 0.744. Pairwise comparisons demonstrated significant differences between all SGS variants except SGS(first HR) vs SGS(mean HR), with the largest AUROC observed for SGS(lowest HR) and the smallest for SGS(highest HR) (Table 4).

#### 3.2.4 Reclassification flow

Reclassification across the SGS dichotomous threshold was common, with substantial movement of cases between low-risk and high-risk categories depending on the choice of HR value employed (Figure 1).

**Figure 1.**
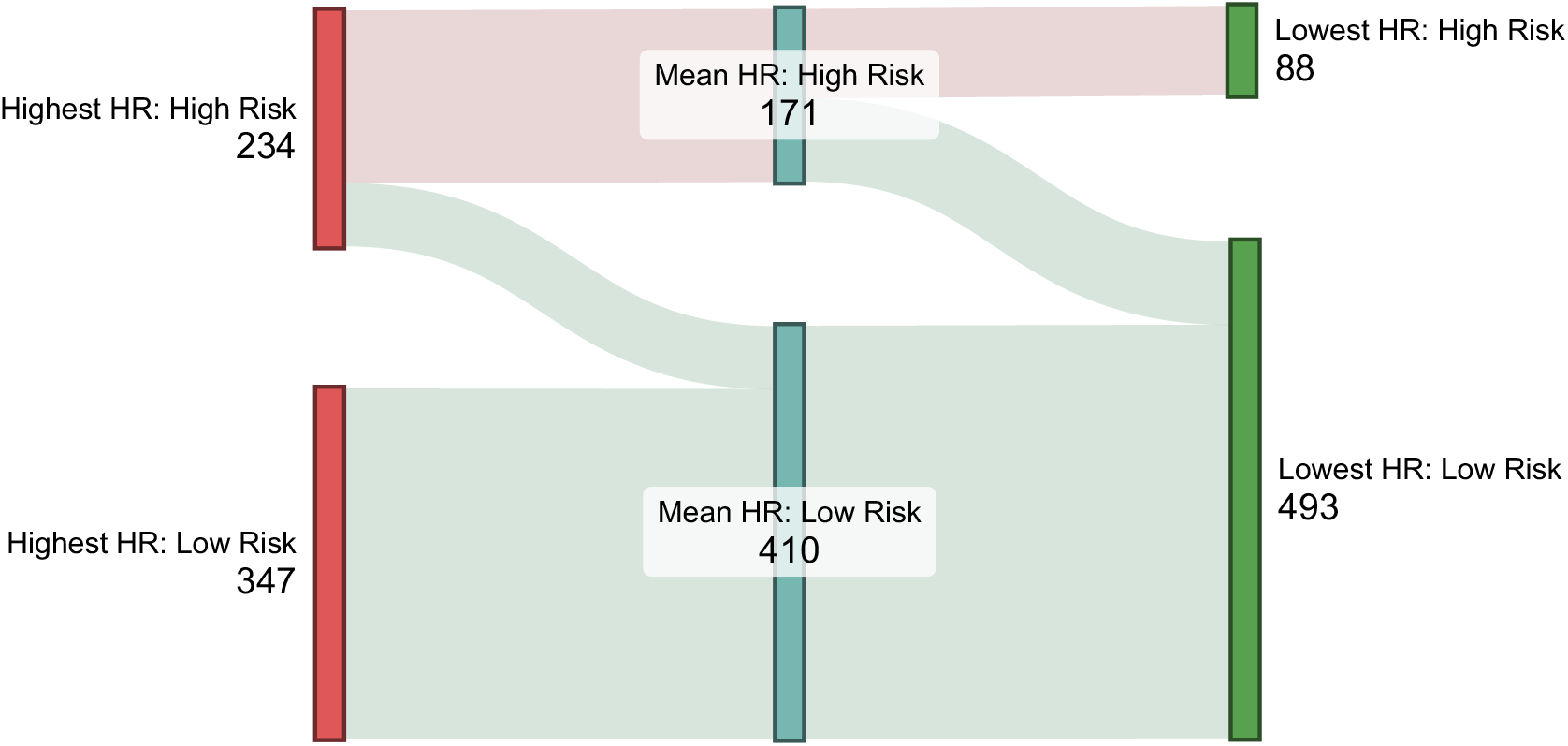
Reclassification flow between SGS(highest HR) -> SGS(mean HR) -> SGS(lowest HR).

The greatest difference was observed between SGS(lowest HR) and SGS(highest HR), with high-risk classification proportions differing by 25 percentage points (15% vs 40% respectively). Within a diagnostic pathway incorporating PERC +/-XDP testing for low-risk patients, use of SGS(lowest HR) would have reduced CTPA utilisation by 11 percentage points relative to SGS(highest HR), 9 percentage points relative to SGS(first HR), and by 7 percentage points relative to SGS(mean HR) (Table 4).

Among the patients reclassified from high-risk to low-risk by SGS(lowest-HR), no PE-positive cases bypassed downstream PERC and/or XDP testing, resulting in no additional missed PEs within the present cohort.

#### 3.2.5 Pathway concordance

Of the 581 CTPA examinations included in the study, 171 (29%) were performed in accordance with the diagnostic pathway, while 410 (71%) were performed despite pathway criteria for imaging not being met. The PE positivity rate among pathway-concordant scans was 17.5% (30/171), compared with 4.9% (20/410) among off-pathway examinations.

## 4.0 Discussion

### 4.1 Principal findings

This study demonstrates that the choice of heart rate value used in calculation of the Simplified Geneva Score substantially influences score output, dichotomous risk classification, diagnostic performance, and downstream imaging utilisation.

The proportion of cases classified as high risk varied from 15% to 40% across SGS implementations, with 146 of 581 episodes (25%) changing classification solely as a consequence of the heart rate value selected.

Reclassification was entirely unidirectional, with lower heart-rate selections resulting exclusively in movement from high-risk to low-risk categories. This consistent directional effect suggests that heart rate selection exerts a real, systematic influence on SGS classification, rather than introducing random variation.

These changes translated into meaningful differences in both diagnostic performance and CTPA utilisation, with the lowest-HR implementation demonstrating the most favourable overall balance of discrimination and imaging efficiency.

### 4.2 Diagnostic performance

The lowest heart rate variant SGS(lowest-HR) demonstrated the highest overall discriminative performance, with its AUROC 0.040 higher than that of SGS(mean-HR), and 0.085 higher than SGS(highest-HR). It also demonstrated substantially higher specificity at 88% (vs 73% and 62% for mean and highest heart rate variants respectively).

This improvement was not confined to a single performance metric. The lowest-HR implementation consistently demonstrated the most favourable AUROC, specificity, NNS, and downstream imaging utilisation. Conversely, the highest-HR implementation consistently performed least favourably across these measures. The mean-HR implementation specified in the original SGS derivation and validation studies was also consistently outperformed by the lowest-HR implementation across each of these metrics.

### 4.3 Diagnostic efficiency vs sensitivity

Although the Lowest-HR implementation demonstrated the highest AUROC and most favourable NNS metrics, these gains were accompanied by reduced sensitivity. This raises concerns about missing an unacceptably large proportion of PEs, but such concern should be tempered by the context in which RSTs such as the SGS should be used, which is as the initial step in a diagnostic pathway, not as a stand-alone tool to fully or adequately exclude PE, in isolation.

Improved RST specificity can reduce unnecessary imaging by more effectively concentrating PEs in the high-risk stratum, while maintaining a sufficiently low PE prevalence within the low-risk group for downstream pathway components to provide the high sensitivity required for overall diagnostic safety.

When embedded within a broader diagnostic pathway incorporating PERC and/or XDP testing, SGS(lowest-HR) produced an NPV of 94.3% (95% CI 93.1-95.7), with a low-risk cohort PE prevalence of only 5.7% (95% CI 4.3 - 6.9), well below the thresholds generally considered acceptable for downstream rule-out strategies (< 15% for PERC and 10-15% for XDP testing alone (6)). Conservatively allowing for up to a doubling of PE prevalence within both SGS risk strata, consistent with the prevalence observed in the SGS derivation and validation cohorts (4,5), the low-risk prevalence associated with SGS(lowest-HR) would increase from 5.7% to 11.4%. This would still remain within the range for which PERC has been validated as a safe rule-out strategy.

Reassuringly, although SGS(lowest-HR) reduced sensitivity at the SGS classification stage, no PE identified within this cohort would have been missed after application of downstream pathway components. This observation supports the contention that improved risk stratification efficiency can be achieved without compromising overall pathway safety.

Accordingly, implementing lowest-HR SGS achieved improved imaging efficiency without incurring clinically unacceptable diagnostic risk, as the reduction in SGS sensitivity remained compatible with accepted downstream rule-out strategies.

### 4.4 Clinical implications

Although the original SGS derivation and validation studies specified use of the mean heart rate when multiple measurements were available, this approach is unlikely to reflect routine bedside practice. In most clinical settings, the highest heart rate or the first measured observed heart rate is more likely to be used when calculating the SGS.

Notably, the highest-HR implementation, which may most closely reflect real-world clinician behaviour, demonstrated the poorest discriminative performance and resulted in both the highest proportion of cases being classified as high risk, and the greatest number of negative CTPA examinations.

From a practical perspective, the differences observed were clinically meaningful. To diagnose 100 pulmonary emboli, use of SGS(lowest-HR) would require approximately 58 fewer CTPA scans than SGS(mean-HR), and approximately 102 fewer than SGS(highest-HR). Given the cost, radiation exposure, contrast exposure, and ED resource utilisation associated with CTPA, such differences are unlikely to be trivial.

These findings were observed in a cohort with a PE prevalence of only 8.6%, despite all patients proceeding to CTPA. Interestingly, in our cohort, pathway-concordant CTPA examinations demonstrated a PE positivity rate of 17.5%, compared with only 4.9% among examinations performed outside pathway criteria. This suggests that departures from pathway-based risk stratification contributed substantially to low-yield imaging and may partially explain the low overall PE prevalence observed within the cohort. Notably, although only 29% of scans were pathway-concordant, these examinations accounted for 60% of all diagnosed pulmonary emboli.

These findings highlight the importance of explicitly defining heart-rate selection methodology in both clinical practice and future studies evaluating RST performance. Furthermore, the findings suggest that deviations from pathway-based investigation contribute substantially to low-yield imaging and may represent a major determinant of overall CTPA utilisation.

### 4.5 Limitations

This study was conducted retrospectively at a single centre, potentially limiting generalisability to other healthcare settings and patient populations. Only patients undergoing CTPA for investigation of suspected PE were included, introducing spectrum bias, and therefore the cohort may not fully reflect the broader population in whom the SGS is applied. Consequently, absolute estimates of PPV, NPV, and NNS may not be directly applicable to unselected ED populations with suspected PE. Additionally, predictive values and NNS estimates are prevalence-dependent and may differ in populations with substantially different underlying rates of PE.

Although consecutive cases were reviewed and only a small number excluded due to incomplete data, retrospective extraction of SGS components remains dependent on the accuracy and completeness of clinical documentation. Furthermore, this study evaluated only alternative heart-rate implementations of the SGS, and the findings may not be directly applicable to other PE risk stratification tools.

Finally, while the lowest-HR implementation demonstrated favourable performance characteristics within this cohort, external validation in independent prospective datasets would be required before any modification to established SGS methodology could be recommended.

## 5.0 Conclusions

The choice of heart rate value used in SGS calculation materially influences score output, risk classification, diagnostic performance, and downstream imaging utilisation in clinically meaningful ways. In this cohort, the lowest-HR implementation demonstrated the most favourable balance of risk discrimination and imaging efficiency, maintaining diagnostic safety when used as part of an established pathway, while the highest-HR implementation demonstrated the least favourable performance. Given the substantial impact of heart-rate selection on SGS behaviour, this implementation detail should be explicitly considered in both clinical practice and future research. These findings provide a rationale for prospective multicentre evaluation to confirm external validity and quantify the impact of heart rate selection on imaging utilisation across diverse clinical settings.

## Data Availability

All de-identified data produced in the present study can be made available upon reasonable request to the author, in consultation with the ACT HREC.

## 6.0 Declarations

### 6.2 AI use disclosure

A generative AI tool (Microsoft Copilot, a GPT-5 based model) was used for language editing, manuscript structuring assistance, and proofreading during final preparation of the manuscript, only. All study design, data extraction, statistical analysis, interpretation of results, and scientific conclusions were performed by the principal investigator, and final manuscript content remained the responsibility of the author.

